# Impact of GLP-1 Receptor Agonist Therapy on Atrial Fibrillation Recurrence After Catheter Ablation in Obese Patients: A Real-World Data Analysis

**DOI:** 10.1101/2025.05.29.25328594

**Authors:** Sandrine Venier, Pascal Defaye, Lisa Lochon, Rémi Benali, Arnaud Bisson, Adrien Carabelli, Youssou Diouf, Peggy Jacon, Laurent Fauchier

**Author notes:** Please address all correspondence to: Dr. Sandrine Venier, Mailing address: Department of Cardiology, University Hospital of Grenoble Centre Alpes France, CS10217, 38043 Grenoble Cedex 9, France.

## Abstract

**Background:** GLP-1 receptor agonists (GLP-1RAs), initially developed for glycemic control in type 2 diabetes, have shown cardiometabolic benefits including weight loss, improved endothelial function, and reduced inflammation. Recent data suggest potential anti-arrhythmic effects via modulation of atrial substrate and autonomic tone. Their impact in obese, non-diabetic patients remains underexplored. This study examines whether GLP-1RA use is associated with reduced AF recurrence after catheter ablation in obese patients, using real-world data from a large multicenter database.

**Methods:** We conducted a retrospective cohort study using the TriNetX research network, which contains de-identified electronic health records from more than 100 million patients. Adult patients (age ≥18 years) with obesity (BMI >30 kg/m^2^) who underwent AF ablation between January 2005 and January 2025 were eligible. The cohort was divided into GLP-1RA users (n = 2,867) and non-users (n = 2,867), with 1:1 propensity score matching performed across 82 clinical and demographic variables including age, sex, race, AF subtype, cardiovascular comorbidities, and baseline medications.

**Results:** During a median follow-up of 1.6 years (IQR: 2.6): AF recurrence was significantly lower in GLP-1RA users (7.43% vs. 8.40%, HR 0.843, 95%CI 0.780-0.911, p<0.0001) Progression to permanent AF occurred less frequently in GLP-1RA users (3.15% vs. 4.35%, HR 0.743, 95%CI 0.610–0.905, p=0.003). Risk of all-cause mortality was lower in the GLP-1RA group (HR 0.700, 95%CI 0.553–0.887, p=0.003) HF hospitalization (HR 0.819, 95%CI 0.722-0.929, p=0.002) and cardiovascular hospitalizations (HR 0.856, 95%CI 0.773-0.947, p=0.003) were also significantly lower with GLP-1RA use. No significant difference was found for redo ablation.

**Conclusion:** In a large real-world cohort of obese patients undergoing catheter ablation for AF, GLP-1RA therapy was associated with lower risks of AF recurrence, progression to permanent AF, cardiovascular hospitalizations, and mortality.

## Introduction

Atrial fibrillation (AF) is the most common sustained cardiac arrhythmia, affecting an estimated 50 million people worldwide and contributing significantly to stroke, heart failure, and increased mortality. As obesity becomes increasingly prevalent, it has emerged as a major risk factor for both the development and recurrence of AF (1). The interplay between excess adiposity, atrial remodeling and systemic inflammation highlights the importance of addressing weight and metabolic health when managing AF. (2,3)

Catheter ablation is an effective rhythm control strategy in AF, offering superior outcomes compared to antiarrhythmic drug (AAD) therapy in selected patients. However, recurrence following ablation remains a frequent challenge, especially in those with persistent modifiable risk factors such as obesity (4,5). Current guidelines emphasize the importance of lifestyle interventions, such as weight loss, in reducing AF burden and recurrence, particularly in patients with a body mass index (BMI) greater than 27 kg/m^2^ (6).

Glucagon-like peptide-1 receptor agonists (GLP-1RAs) have gained prominence for two reasons: their glucose-lowering effects and their ability to induce significant weight loss (7,8). Initially developed for glycemic control in type 2 diabetes, GLP-1RAs have also demonstrated cardiometabolic benefits, improved endothelial function and reduced inflammation (9,10). Recent data also suggest potential anti-arrhythmic effects, particularly through modulation of atrial substrate and autonomic tone (11,12). While a recent study by Satti et al. (13) found no reduction in AF recurrence following GLP-1RA therapy in a diabetic population, the specific impact of GLP-1RAs in obese, non-diabetic patients remains under- explored. These effects make GLP-1RAs a potentially valuable adjunct in the management of AF, particularly in obese individuals (14).

In this study, we aim to investigate whether GLP-1RA use at the time of catheter ablation is associated with reduced AF recurrence and improved cardiovascular outcomes specifically in obese patients. Our aim in isolating this high-risk population is to evaluate whether metabolic modulation through GLP-1RA therapy can result in significant arrhythmia-related clinical benefits following ablation.

## Methods

We conducted a retrospective cohort study using the TriNetX research network, which contains de-identified electronic health records from more than 100 million patients. Adult patients (age ≥18 years) with obesity (BMI >30 kg/m^2^) who underwent AF ablation between January 2005 and January 2025 were eligible. The cohort was divided into GLP-1RA users (n = 2,867) and non-users (n = 2,867), with 1:1 propensity score matching performed across 82 clinical and demographic variables including age, sex, race, AF subtype, cardiovascular comorbidities, and baseline medications.

### Data Source

We utilized the TriNetX Global Collaborative Network, a federated health research database composed of anonymized electronic medical records from 143 healthcare organizations. TriNetX aggregates information on diagnoses, procedures, medications, laboratory values, and demographics, supporting patient-level analytics across a wide and diverse population.

### Patient Population

We included adult patients (aged 18–90 years) with documented obesity (BMI ≥30 kg/m^2^ or ICD-10 diagnosis codes for obesity) who underwent AF ablation between January 2005 and January 2025. Ablation procedures were identified using CPT code 93656. Patients were categorized into two cohorts based on GLP-1RA use: those who had a prescription for GLP-1RA within one year prior to or up to six months after the index ablation (GLP-1RA group), and those without any GLP-1RA exposure during the same timeframe (control group).

### Exposure Definition

GLP-1RA use included any prescription of semaglutide, liraglutide, dulaglutide, exenatide, lixisenatide, or tirzepatide. Only patients with concurrent evidence of obesity (laboratory BMI ≥30 kg/m^2^ or ICD-10 codes E66.0-E66.9) were included.

### Index Event and Follow-up

The index event was defined as the date of the first recorded catheter ablation procedure for AF. Outcomes were assessed from one day post-index and continued up to six years post-ablation (2190 days). Patients with index events more than 20 years prior to analysis were excluded.

### Study Outcomes

The primary outcome was recurrence of AF, defined as hospitalization or outpatient visit coded for AF. Secondary outcomes included progression to permanent AF, all-cause mortality, hospitalization for heart failure, cardiovascular-related hospitalizations, repeat ablation procedures, AV node ablation, and a composite of cardioversion or amiodarone initiation.

### Statistical Analysis

Cohorts were matched 1:1 using propensity score matching (PSM) with a nearest-neighbor algorithm and a caliper width of 0.1 pooled standard deviations. PSM was conducted across 82 variables, including demographics, comorbidities, AF subtype, laboratory results (e.g. BMI and HbA1c) and medication usage. Covariate balance was assessed using standardized mean differences (SMDs). SMDs <0.1 indicate good balance. Outcome incidence was analyzed using both risk and survival metrics. Hazard ratios (HR) with 95% confidence intervals (CI) were calculated, and Kaplan-Meier curves were generated to compare time-to-event data.

### Subgroup Analyses

Outcomes were further analyzed by GLP-1RA type, comparing semaglutide versus other agents. Results were stratified to evaluate potential differential effects based on the specific medication used.

### Ethics

All data were de-identified in accordance with HIPAA standards and the analysis was conducted in compliance with institutional and regulatory guidelines for retrospective, anonymized research.

## Results

The overall cohort selection flow is presented in Figure 1. Following the application of inclusion and exclusion criteria, a total of 5,734 obese patients who underwent atrial fibrillation (AF) ablation were included in the analysis. Semaglutide was the most common GLP-1RA used in the cohort. After performing 1:1 propensity score matching, two well-balanced cohorts of 2,867 patients each were established: one receiving GLP-1RA therapy and the other not. As shown in Table 1, the matching process achieved covariate balance across demographic, clinical, and laboratory characteristics, with standardized mean differences below 0.1 for all variables. Supplemental Figure 1 confirms the successful propensity score matching through density plots pre- and post-matching.

**Figure 1.**
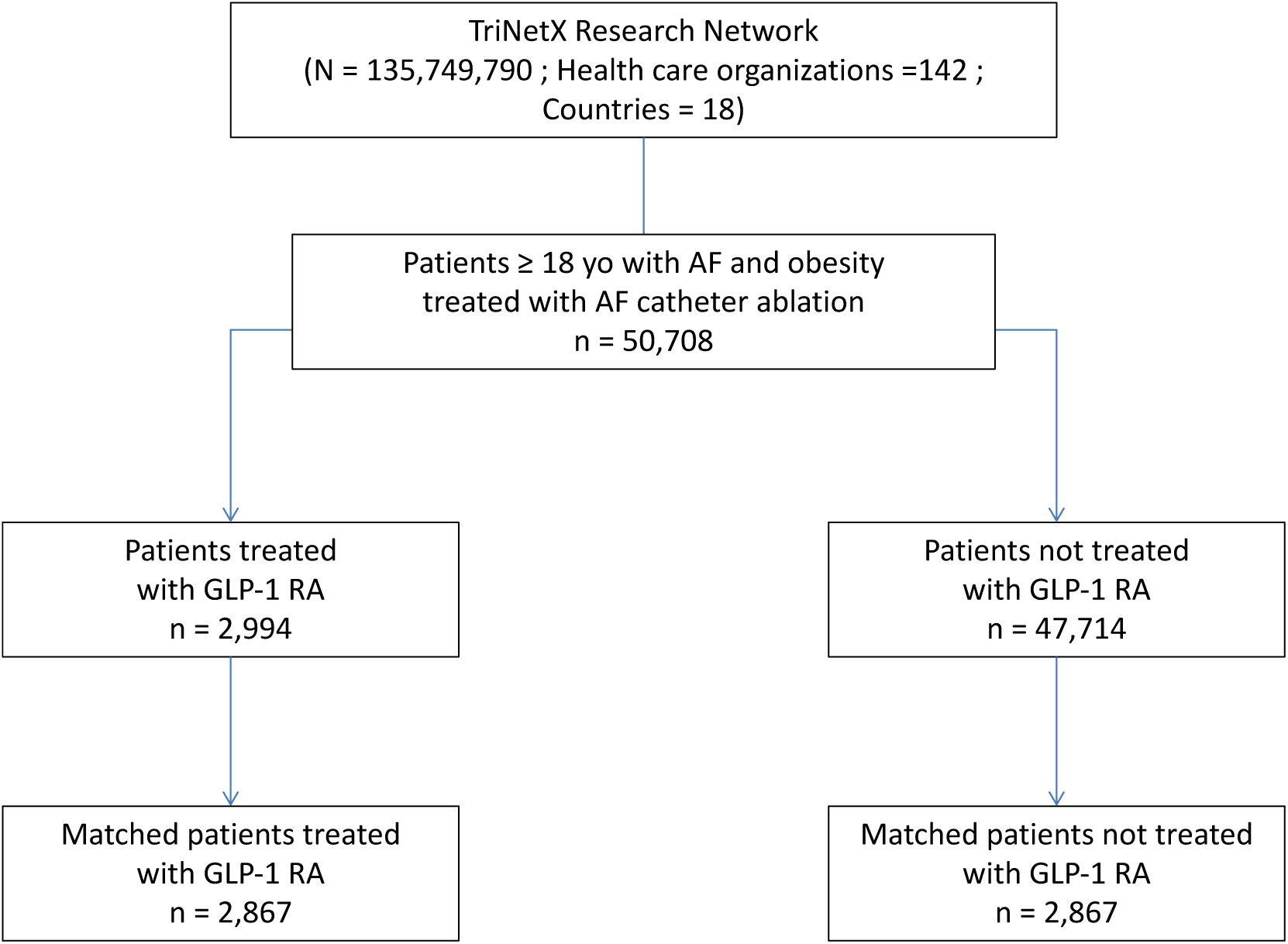
Flow chart of patients in the study

**Table 1.** Baseline characteristics of patients before and after propensity score matching.

|  | Before propensity-score matching |  |  | After propensity-score matching |  |  |
| --- | --- | --- | --- | --- | --- | --- |
|  | AF ablation,<br>GLP-1RA<br>(n = 2994) | AF ablation,<br>no GLP-1RA<br>(n = 47714) | Std<br>diff.<br>(%) | AF ablation,<br>GLP-1RA<br>(n = 2867) | AF ablation,<br>no GLP-1RA<br>(n = 2867) | Std<br>diff.<br>(%) |
| *Demographics* |  |  |  |  |  |  |
| **Age (years), mean±SD** | 63.5 +/- 9.3 | 64.5 +/- 10.3 | 10.6 | 63.6 +/- 9.3 | 64.1 +/- 10.0 | 4.8 |
| **Men, n (%)** | 1830 (61.1%) | 31406 (65.8%) | 9.8 | 1752 (61.1%) | 1729 (60.3%) | 1.6 |
| **White, n (%)** | 2436 (81.4%) | 40245 (84.3%) | 7.9 | 2326 (81.1%) | 2340 (81.6%) | 1.3 |
| **Black or African American, n (%)** | 324 (10.8%) | 3370 (7.1%) | 13.2 | 314 (11%) | 304 (10.6%) | 1.1 |
| **Asian, n (%)** | 37 (1.2%) | 549 (1.2%) | 0.8 | 36 (1.3%) | 41 (1.4%) | 1.5 |
| **Hispanic or Latino, n (%)** | 130 (4.3%) | 1298 (2.7%) | 8.8 | 118 (4.1%) | 112 (3.9%) | 1.1 |
| **Socioeconomic and psychosocial circumstances, n (%)** | 120 (4%) | 843 (1.8%) | 13.4 | 111 (3.9%) | 107 (3.7%) | 0.7 |
| *Risk factors* |  |  |  |  |  |  |
| **Hypertension, n (%)** | 2578 (86.1%) | 33885 (71%) | 37.4 | 2457 (85.7%) | 2473 (86.3%) | 1.6 |
| **Diabetes mellitus, n (%)** | 2326 (77.7%) | 11673 (24.5%) | 125.8 | 2199 (76.7%) | 2247 (78.4%) | 4 |
| **Smoker, n (%)** | 870 (29.1%) | 12587 (26.4%) | 6 | 833 (29.1%) | 854 (29.8%) | 1.6 |
| **Dyslipidaemia, n (%)** | 2470 (82.5%) | 29264 (61.3%) | 48.5 | 2350 (82%) | 2343 (81.7%) | 0.6 |
| **Alcohol related diagnoses, n (%)** | 88 (2.9%) | 1421 (3%) | 0.2 | 83 (2.9%) | 91 (3.2%) | 1.6 |
| *Cardiovascular comorbidities* |  |  |  |  |  |  |
| **Heart failure, n (%)** | 1418 (47.4%) | 16512 (34.6%) | 26.2 | 1366 (47.6%) | 1409 (49.1%) | 3 |
| **Coronary artery disease, n (%)** | 1502 (50.2%) | 17552 (36.8%) | 27.2 | 1431 (49.9%) | 1447 (50.5%) | 1.1 |
| **Myocardial infarction, n (%)** | 298 (10%) | 3131 (6.6%) | 12.3 | 286 (10%) | 280 (9.8%) | 0.7 |
| **Ischemic stroke, n (%)** | 173 (5.8%) | 1931 (4%) | 8 | 159 (5.5%) | 167 (5.8%) | 1.2 |
| **Intracranial hemorrhage, n (%)** | 19 (0.6%) | 102 (0.2%) | 6.5 | 19 (0.7%) | 12 (0.4%) | 3.3 |
| **Valve disease, n (%)** | 651 (21.7%) | 9820 (20.6%) | 2.8 | 634 (22.1%) | 632 (22%) | 0.2 |
| **Pulmonary embolism/DVT, n (%)** | 75 (2.5%) | 956 (2%) | 3.4 | 70 (2.4%) | 79 (2.8%) | 2 |
| **Paroxysmal AF, n (%)** | 2451 (81.9%) | 34156 (71.6%) | 24.5 | 2333 (81.4%) | 2339 (81.6%) | 0.5 |
| *Non-cardiovascular comorbidities* |  |  |  |  |  |  |
| **Kidney disease, n (%)** | 997 (33.3%) | 9323 (19.5%) | 31.6 | 950 (33.1%) | 961 (33.5%) | 0.8 |
| **COPD, n (%)** | 416 (13.9%) | 4587 (9.6%) | 13.3 | 394 (13.7%) | 358 (12.5%) | 3.7 |
| **Sleep apnoea syndrome, n (%)** | 1945 (65%) | 19950 (41.8%) | 47.7 | 1836 (64%) | 1853 (64.6%) | 1.2 |
| **Peripheral vascular disease, n (%)** | 232 (7.7%) | 2162 (4.5%) | 13.4 | 223 (7.8%) | 249 (8.7%) | 3.3 |
| **Previous cancer, n (%)** | 845 (28.2%) | 8947 (18.8%) | 22.5 | 799 (27.9%) | 822 (28.7%) | 1.8 |
| **Cognitive impairment, n (%)** | 10 (0.3%) | 52 (0.1%) | 4.8 | 10 (0.3%) | 10 (0.3%) | 0 |
| *Laboratory tests and examinations* |  |  |  |  |  |  |
| **Heart rate, mean±SD** | 75.2 +/- 16.6 | 71.4 +/- 17.9 | 21.8 | 75.0 +/- 16.6 | 74.2 +/- 18.3 | 4.9 |
| **Body mass index (kg/m2), mean±SD** | 37.6 +/- 6.6 | 34.6 +/- 5.5 | 49.7 | 37.5 +/- 6.6 | 37.0 +/- 6.4 | 7.7 |
| **Total cholesterol (mg/dL), mean±SD** | 147.7 +/- 40.2 | 158.4 +/- 43.2 | 25.6 | 148.2 +/- 40.6 | 148.8 +/- 41.3 | 1.5 |
| **LDL cholesterol (mg/dL), mean±SD** | 75.9 +/- 32.9 | 87.7 +/- 34.3 | 35.3 | 76.4 +/- 33.3 | 77.9 +/- 33.8 | 4.6 |
| **HDL cholesterol (mg/dL), mean±SD** | 41.0 +/- 17.0 | 42.5 +/- 19.2 | 8.2 | 41.1 +/- 17.1 | 39.9 +/- 18.0 | 6.6 |
| **Triglyceride (mg/dL), mean±SD** | 151.7 +/- 94.4 | 130.2 +/- 83.4 | 24.1 | 150.9 +/- 95.0 | 147.8 +/- 102.5 | 3.1 |
| **Estimated GFR (MDRD, ml/min), mean±SD** | 68.4 +/- 23.5 | 70.3 +/- 21.3 | 8.5 | 68.3 +/- 23.5 | 67.5 +/- 23.4 | 3.6 |
| **HbA1c <6%, n (%)** | 1266 (42.3%) | 14007 (29.4%) | 27.2 | 1205 (42%) | 1274 (44.4%) | 4.9 |
| **HbA1c ≥6%, n (%)** | 1528 (51%) | 4777 (10%) | 99.5 | 1411 (49.2%) | 1391 (48.5%) | 1.4 |
| **Hemoglobin (g/dl), mean±SD** | 13.5 +/- 1.8 | 13.9 +/- 1.8 | 17.6 | 13.5 +/- 1.8 | 13.5 +/- 1.9 | 1 |
| **BNP, ng/L, mean±SD** | 342.7 +/- 696.6 | 427.4 +/- 1050.8 | 9.5 | 348.5 +/- 711.5 | 383.1 +/- 710.3 | 4.9 |
| **NT-proBNP, ng/L, mean±SD** | 1507.2 +/- 3697.4 | 2138.9 +/- 4406.4 | 15.5 | 1554.5 +/- 3778.6 | 1639.5 +/- 3510.9 | 2.3 |
| **LVEF <40%, n (%)** | 161 (5.4%) | 2236 (4.7%) | 3.2 | 158 (5.5%) | 166 (5.8%) | 1.2 |
| **LA diameter (anterior-posterior, mm), mean±SD** | 36.7 +/- 14.2 | 33.5 +/- 17.0 | 19.9 | 36.5 +/- 14.4 | 35.8 +/- 15.5 | 5 |
| **LV end-diastolic diameter (cm), mean±SD** | 4.9 +/- 0.7 | 4.8 +/- 0.7 | 7.3 | 4.9 +/- 0.7 | 4.8 +/- 0.9 | 4.2 |
| *Medications* |  |  |  |  |  |  |
| **Beta Blockers, n (%)** | 2497 (83.4%) | 36162 (75.8%) | 19 | 2377 (82.9%) | 2404 (83.9%) | 2.5 |
| **Calcium Channel Blockers, n (%)** | 1713 (57.2%) | 22453 (47.1%) | 20.4 | 1633 (57%) | 1679 (58.6%) | 3.2 |
| **ACE Inhibitors, n (%)** | 997 (33.3%) | 11982 (25.1%) | 18.1 | 936 (32.6%) | 913 (31.8%) | 1.7 |
| **Angiotensin II Inhibitors, n (%)** | 1374 (45.9%) | 13305 (27.9%) | 38 | 1310 (45.7%) | 1328 (46.3%) | 1.3 |
| **ARNI, n (%)** | 330 (11%) | 2272 (4.8%) | 23.4 | 307 (10.7%) | 284 (9.9%) | 2.6 |
| **MRA, n (%)** | 708 (23.6%) | 5948 (12.5%) | 29.4 | 668 (23.3%) | 667 (23.3%) | 0.1 |
| **Digitalis glycosides, n (%)** | 338 (11.3%) | 4408 (9.2%) | 6.8 | 322 (11.2%) | 354 (12.3%) | 3.5 |
| **Diuretics, n (%)** | 2216 (74%) | 27325 (57.3%) | 35.8 | 2123 (74%) | 2167 (75.6%) | 3.5 |
| **Lipid lowering drugs, n (%)** | 2414 (80.6%) | 26000 (54.5%) | 58.1 | 2292 (79.9%) | 2323 (81%) | 2.7 |
| **Antiplatelet therapy, n (%)** | 1467 (49%) | 19234 (40.3%) | 17.5 | 1407 (49.1%) | 1431 (49.9%) | 1.7 |
| **Anticoagulant, n (%)** | 1854 (61.9%) | 28715 (60.2%) | 3.6 | 1770 (61.7%) | 1798 (62.7%) | 2 |
| **Amiodarone, n (%)** | 1157 (38.6%) | 14748 (30.9%) | 16.3 | 1106 (38.6%) | 1127 (39.3%) | 1.5 |
| **Flecainide, n (%)** | 396 (13.2%) | 7350 (15.4%) | 6.2 | 375 (13.1%) | 392 (13.7%) | 1.7 |
| **Propafenone, n (%)** | 57 (1.9%) | 1627 (3.4%) | 9.4 | 55 (1.9%) | 53 (1.8%) | 0.5 |
| **Sotalol, n (%)** | 280 (9.4%) | 4642 (9.7%) | 1.3 | 272 (9.5%) | 256 (8.9%) | 1.9 |
| **Dronedarone, n (%)** | 170 (5.7%) | 3019 (6.3%) | 2.7 | 162 (5.7%) | 164 (5.7%) | 0.3 |

\* **GLP-1RA**= Glucagon-Like Peptide-1 Receptor Agonist **SD**= Standard Deviation **GFR**= Glomerular Filtration Rate **NT-BNP**= N-terminal pro-B-type Natriuretic Peptide **LVEF**= Left Ventricular Ejection Fraction **LA**= Left Atrium **LV**= Left ventricle **ACE**= Angiotensin-Converting Enzyme **ARNI**= Angiotensin Receptor–Neprilysin Inhibitor **MRA**= Mineralocorticoid Receptor Antagonist

Kaplan–Meier survival curves for major clinical outcomes are provided in Figure 2. During a median follow-up period of 1.6 years (interquartile range 2.6), GLP-1RA users experienced a significantly lower rate of AF recurrence requiring hospitalization or outpatient visit, with an incidence of 7.43% compared to 8.40% in the non-GLP-1RA group (HR 0.843, 95% CI: 0.780–0.911; p < 0.0001), (Figure 2A).

**Figure 2.**
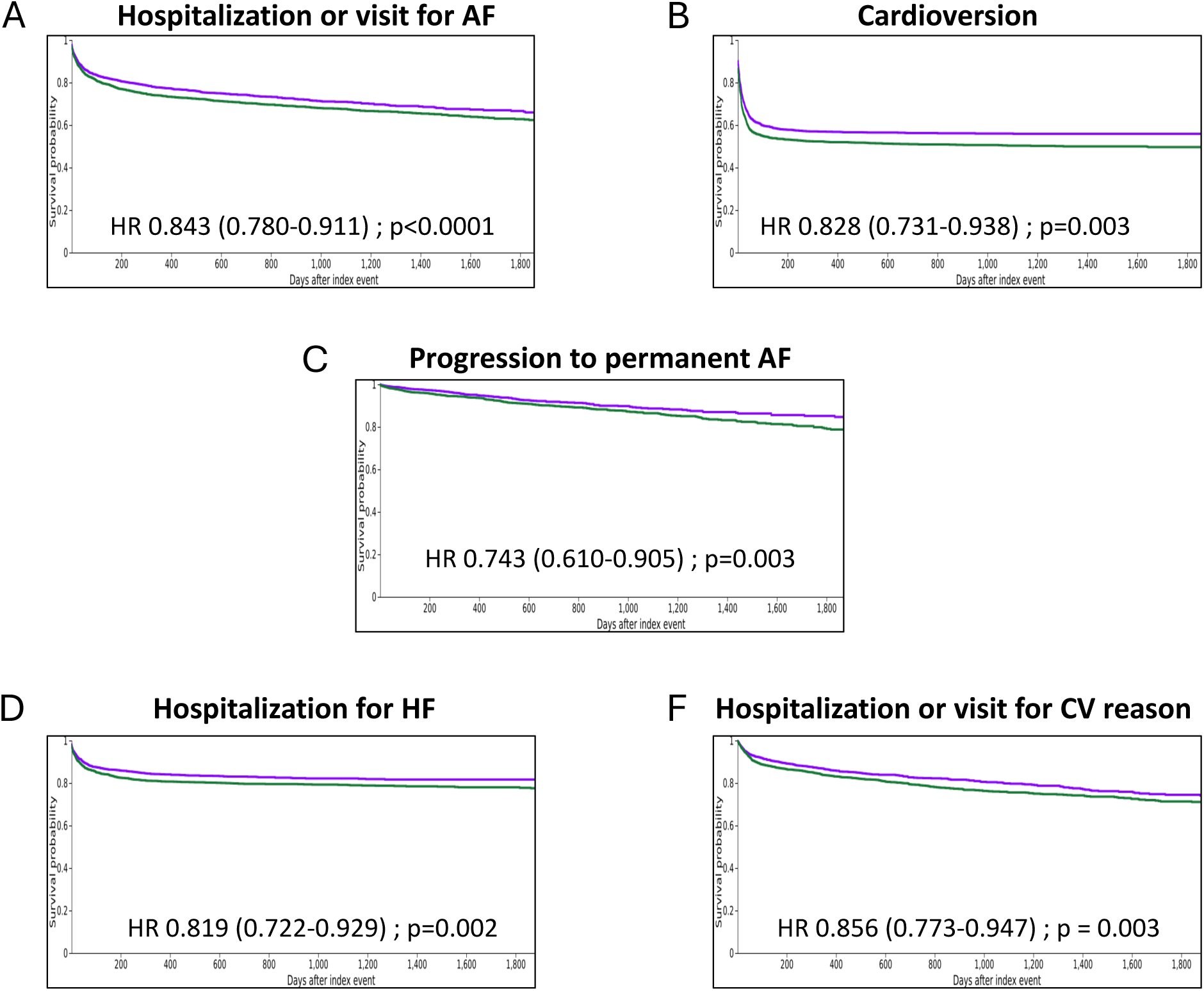
Kaplan–Meier analysis with event-free probabilities in patients with obesity after AF ablation for A) Hospitalisation or visit for AF, B) Cardioversion, C) Progression to permanent AF, D) Hospitalisation for HF, and E) Hospitalisation or visit for CV reason

Regarding other clinical outcomes, the GLP-1RA group showed a significantly lower risk of progression to permanent AF, occurring in 3.15% of patients compared to 4.35% in the control group (HR 0.743, 95% CI: 0.610–0.905; p = 0.003), with results shown in Figure 2C. Risk of all-cause mortality was also significantly lower in the GLP-1RA group (HR 0.700, 95% CI: 0.553–0.887; p = 0.003). Hospitalizations for heart failure and cardiovascular-related causes were both less frequent among GLP-1RA users, with hazard ratios of 0.819 (95%CI 0.722-0.929, p=0.002) and 0.856 (95%CI 0.773-0.947, p=0.003), respectively (Figures 2D and 2E). Additionally, risk of all-cause hospitalization was significantly lower in the GLP- 1RA group (HR 0.911, 95% CI: 0.837–0.992; p = 0.03).

No significant differences were observed in the rates of repeat AF ablation (HR 1.043; p=0.6), AV node ablation (HR 0.977; p=0.92), or the initiation of amiodarone therapy (HR 0.931; p=0.48). However, the composite endpoint encompassing cardioversion, amiodarone initiation, redo ablation, or AV node ablation showed a significant lower risk in the GLP-1RA group (HR 0.879, 95% CI: 0.811–0.953; p=0.002). These results are summarised in Table 2, which provides an overview of the number of events, yearly rates, hazard ratios and corresponding p-values.

**Table 2.**
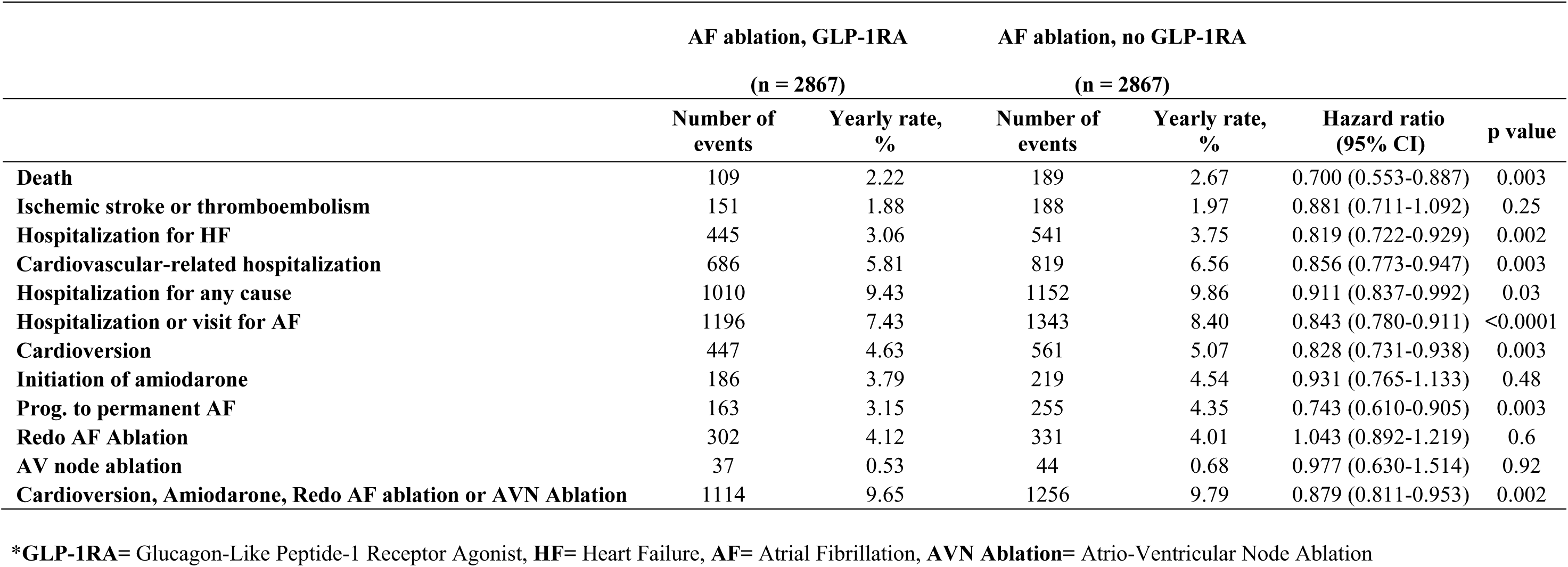
Clinical outcomes during FU in the matched population. (FU 2.1 ± 1.8 years, median 1.6, IQR 2.6)

Subgroup analyses provided additional insights. Patients treated with semaglutide appeared to derive relatively similar clinical benefit than those on other GLP-1RAs since none of the interactions analyses was statistically significant in the separate analyses of risk for the different clinical outcomes comparing semaglutide vs no GLP-1RA and GLP-1RA use (excluding semaglutide) vs no GLP-1RAs. However, all-cause mortality was significantly lower (HR 0.547, 95% CI: 0.343–0.872), and AF recurrence was also significantly lower (HR 0.85, 95% CI: 0.770–0.938) with semaglutide use vs no GLP-1RA. A similar result was observed for heart failure hospitalization (HR 0.809, 95% CI: 0.685–0.956), while other comparisons between GLP-1RA vs no GLP-1RA did not reach statistical significance. Further interaction details are available in Table 3.

**Table 3.**
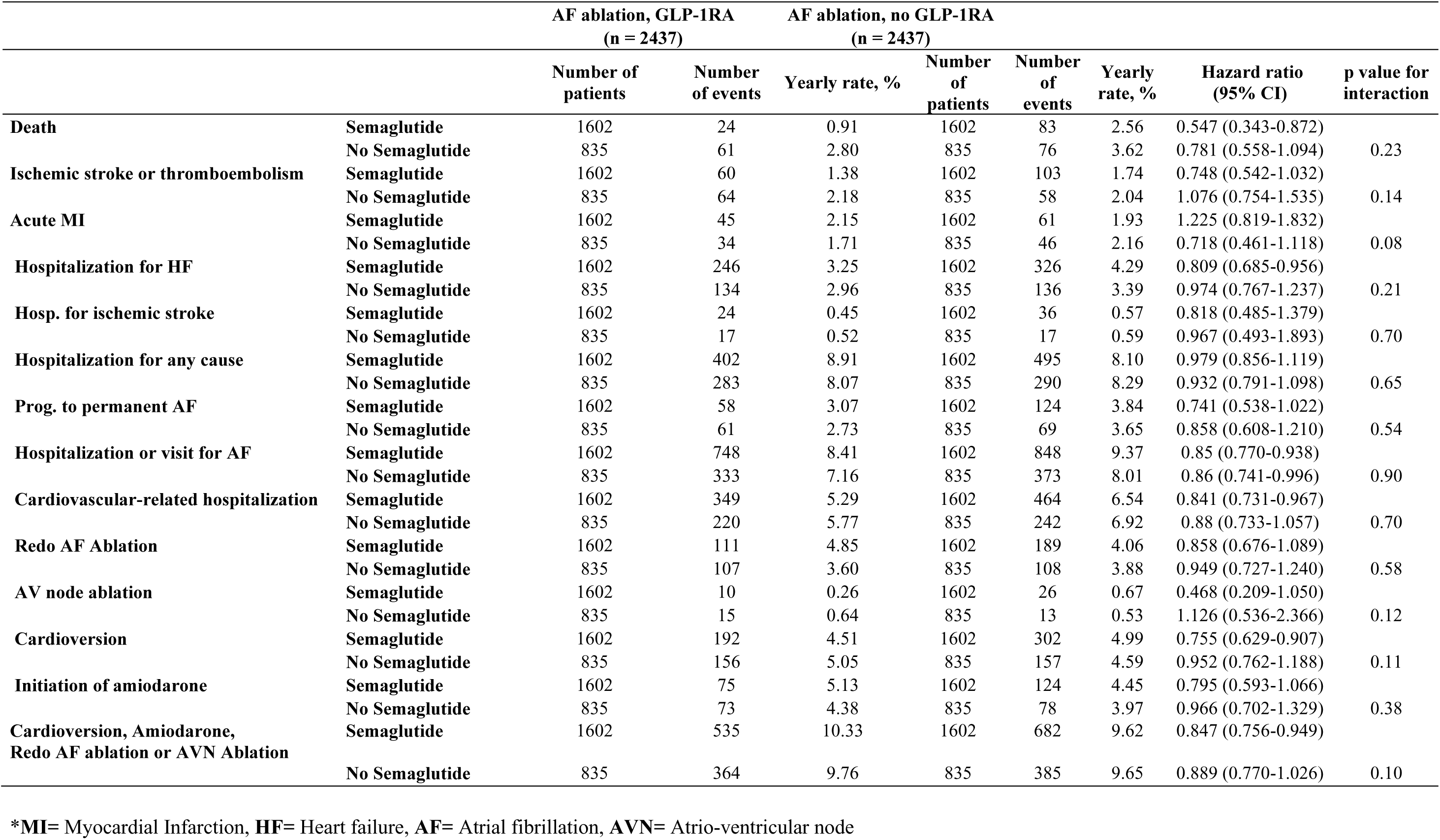
Interaction analysis for clinical outcomes with GLP-1 RA use vs no use according to Semaglutide or another GLP-1RA use.

## Discussion

In this large population-based study of obese patients undergoing AF ablation, GLP-1RA therapy was significantly associated with lower risks of AF recurrence, progression to permanent AF, cardiovascular hospitalizations, and all-cause mortality. To our knowledge, this is the first real-world analysis to examine the impact of GLP-1RA therapy on post-ablation outcomes specifically in an obese cohort.

Our findings contrast with a recent work by Satti et al. (13) which found no significant association between GLP-1RA use and AF recurrence in a broader diabetic population (in whom 35% only had BMI>30). The favorable outcomes observed in our study may be due to our exclusive focus on obese patients, who are particularly susceptible AF recurrence due to atrial remodeling and systemic inflammation linked to adiposity. This supports the hypothesis that weight loss and metabolic control through GLP-1RAs may exert protective effects in AF management.

Our results also align with observational data and meta-analyses suggesting a potential direct or indirect anti-arrhythmic benefit of GLP-1RAs. For example, Shi et al. conducted a network meta-analysis which reported a lower incidence of atrial fibrillation (AF) in diabetic patients treated with GLP-1RAs compared to those treated with other glucose-lowering therapies (14). Furthermore, GLP-1 receptors are expressed in atrial myocardium, and preclinical models have shown that GLP-1RA treatment can prevent atrial remodeling and reduce AF susceptibility (15). Weight loss associated with GLP-1 RAs may contribute to their potential protective effect against AF in obese patients, though this is presented as a general consideration rather than a proven mechanism (16). The overall evidence suggests that GLP- 1RA do not increase AF risk in patients with type 2 diabetes, with most studies showing neutral or potentially protective effects (17,18). One study specifically examined obese patients and found a significant lower risk of AF (27% lower risk in AF incidence) with GLP- 1 receptor agonist use (19).

These findings have important clinical implications. With obesity affecting over 35% of adults in the US and being known to impact AF burden, GLP-1RAs could be used as part of dual-purpose therapy for rhythm control. While lifestyle-induced weight loss is a Class I recommendation for AF management in patients with BMI >27 kg/m^2^, pharmacologic weight loss using GLP-1RAs may represent an effective adjunctive approach.

Our results suggest that GLP-1RA therapy may offer obese patients additional protection against AF recurrence following catheter ablation, in contrast to findings in diabetic cohorts where such benefits were not observed (13). This highlights the potential importance of obesity-specific mechanisms. The mechanisms underlying these findings may include weight loss-induced atrial remodeling reversal, reduced epicardial fat and inflammation, improved glycemic control, and potential direct electrophysiological effects. GLP-1 receptors are expressed in atrial tissue, and preclinical studies have demonstrated atrial anti-fibrotic and anti-arrhythmic properties of these agents (14). In our study, GLP-1RA as a class were associated with a reduced risk of atrial fibrillation recurrence, but semaglutide did not emerge as significantly more effective than other GLP-1RA. While the subgroup analyses indicated numerically more pronounced reductions in mortality and AF recurrence with semaglutide vs no GLP-1RA use by comparison to other GLP-1RAs among patients with prior AF ablation, the statistical analysis indicated that semaglutide did not confer different results compared to other GLP-1RAs. However, recent findings from a meta-analysis by Cesaro et al. suggest that semaglutide may provide a distinct protective benefit, showing a 17% reduction in new-onset AF, particularly with the oral formulation and in the absence of concomitant SGLT2 inhibitor use (20). Nevertheless, our study lends support to the hypothesis that GLP-1RAs may improve arrhythmic outcomes in obese patients undergoing AF ablation, suggesting that prospective randomized trials on this issue are warranted.

### Limitations

This study has several limitations. First, the retrospective nature of the TriNetX database precludes causal inference. Second, the use of administrative data to define outcomes such as AF recurrence may underestimate true burden of AF. AF recurrence was assessed using several surrogate markers such as hospitalizations and procedures, which may underestimate true recurrence rates based on a systematic analysis of ECG by an adjudication committee, which was not possible considering the structure of the analysis. Third, we could not measure the precise degree of weight loss achieved by GLP-1RA use or duration of therapy in each patient, limiting interpretation of the mechanism behind observed effects. Weight loss trajectories were not available, preventing assessment of whether benefit was mediated by weight loss per se or direct drug effects. We were not able to study whether a dose effect was present for GLP-1RA use and clinical outcomes since dosing were not homogeneously available in the healthcare systems of the research network. Fourth, as with all real-world database studies, residual confounding is possible despite robust propensity score matching. A better holistic management of AF in patient treated with GLP-1RA is possible and may also explain some of the present results.

## Conclusion

In obese patients undergoing AF ablation, GLP-1RA use was associated with improved clinical outcomes, including reduced AF recurrence and mortality. These findings support the potential utility of GLP-1RAs as part of a comprehensive management strategy in high-risk obese AF populations and underscore the need for prospective trials to validate these results.

## Data Availability

To gain access to the data in the TriNetX research network, requests are directed to TriNetX and a data sharing agreement is required.

## Ethics Statement

As the study was conducted retrospectively and patients were not directly involved in its conduct, there was no impact on their care. Ethical approval was not required, as all data were anonymized.

---

**Supplemental Figure 1.** Propensity score density function - Before and after matching

